# Patient Benefit and Risk in Anticancer Drug Development: A Systematic Review of the Ixabepilone Trial Portfolio

**DOI:** 10.1101/19003467

**Authors:** Benjamin Carlisle, James Mattina, Tiger Zheng, Jonathan Kimmelman

## Abstract

**OBJECTIVE:** To describe the patient burden and benefit, and the dynamics of trial success in the development of ixabepilone—a drug that was approved in the US but not in Europe.

**DATA SOURCES:** Trials were captured by searching Embase and MEDLINE on July 27, 2015.

**STUDY SELECTION:** Inclusion: 1) primary trial reports, 2) interventional trials, 3) human subjects, 4) phase 1 to phase 3, 5) trials of ixabepilone in monotherapy or combination therapy of 6) pre-licensure cancer indications. Exclusion: 1) secondary reports, 2) interim results, 3) meta-analyses, 4) retrospective/observational studies, 5) laboratory analyses (*ex vivo* tissues), 6) reviews, 7) letters, editorials, guidelines, interviews, abstract-only and poster presentations.

**DATA EXTRACTION AND SYNTHESIS:** Data were independently double-extracted and differences between coders were reconciled by discussion.

**MAIN OUTCOMES AND MEASURES:** We measured risk using the number of drug-related adverse events that were grade 3 or higher, benefit by objective response rate and trial outcomes by whether studies met their primary endpoint with acceptable safety.

**RESULTS:** We identified 39 publications of ixabepilone monotherapy and 23 primary publications of combination therapy, representing 5615 patients and 1598 patient-years of involvement over 11 years and involving 17 different malignancies. In total, 830 patients receiving ixabepilone experienced objective tumour response (16%, 95% CI 12.5%–20.1%), and 74 died from drug-related toxicites (2.2%, 95% CI 1.6%–2.9%). Responding indications and combinations were identified very quickly; thereafter, the search for additional responding indications or combinations did not lead to labelling additions. A total of 11 “uninformative” trials were found, representing 27% of studies testing efficacy, 208 grade 3–4 events and 226 patient-years of involvement (21% and 26% of the portfolio total, respectively). After the European Medicines Agency rejected ixabepilone for licensing, all further trial activity involving ixabepilone was pursued outside of Europe.

**DISCUSSION:** Risk/benefit for patients who enrolled in trials of non-approved indications of ixabepilone did not improve over the course of the drug’s development. Clinical value was discovered very quickly; however, a large fraction of trials were uninformative.

## Introduction

In cancer, approximately one in twenty drugs put into phase 1 testing is ultimately licensed.^1^ The success rates for clinical approval in the first, second and third indications pursued are 9.0, 8.2 and 6.9%, respectively.^2^ This high rate of failure results in burdens for patients—particularly when drugs have substantial toxicity. It also taxes research systems, and indirectly, healthcare systems. Increasingly, research sponsors, policymakers, and drug developers are seeking ways of reducing burdens and improving the efficiency of developing new drugs.

In a previous investigation of the targeted drug sunitinib, we found that rapid licensure led to a dramatic expansion in activities aimed at extending the sunitinib label to other indications. However, much of this later exploratory activity appeared to be duplicative, and risk/benefit worsened as drug developers attempted to extend sunitinib to other indications.^3^

To better explore the dynamics of risk, benefit and success in cancer drug development, we investigated the development of a non-targeted anticancer drug that was developed around the same time as sunitinib. Ixabepilone is a semisynthetic analog of epothilone B.^4^ Ixabepilone was first licensed in 2007 by the FDA for second-line monotherapy and combination therapy with capecitabine in breast cancer.^5^ The National Comprehensive Cancer Network’s guidelines recommended ixabepilone for treatment of HER 2-negative breast cancer,^6^ and NCCN also made a Category 2B (lower-level evidence) recommendation of second-line ixabepilone monotherapy for recurrent, metastatic or high-risk endometrial carcinoma.^7^ However, ixabepilone was withdrawn from development in the European market in November of 2008 due to a refused licensing application to the European Medicines Agency.^8^

## Methods

Our primary goal was to quantify the total patient risk and benefit associated with the development of ixabepilone as a cancer treatment. We defined a “drug development trial portfolio” as the complete set of published clinical trials in ixabepilone monotherapy or combination therapy for oncology for indications that were not approved at the time of trial launch. Benefit was measured in terms of response to therapy by RECIST criteria, and we used a number of measures of patient risk and burden, including treatment-related death rate and grade 3–4 serious adverse event rate by CTCAE criteria. Our secondary goals were to track the evolution of risk and benefit and the relationship between risk/benefit and funding. Because ixabepilone is licensed for both monotherapy and combination therapy with capecitabine, we included both monotherapy and combination therapy trials in our analysis. Our analysis included only trials in non-approved indications for ixabepilone. These methods have been adapted from our previously published systematic review of sunitinib.^3^

## Literature search

Trials were captured by searching Embase and MEDLINE on July 27, 2015 using variations on the drug terms “ixabepilone,” “Azaepothilone B”, “Ixempra”, “BMS-247550”, “BMS 247550-1” and “aza-epothilone B.” These results were combined with exploded MeSH terms including “randomized controlled trial,” “clinical trials,” and “phase 1” to “phase 3.” The full search protocol is provided in our appendix.

Publications were screened for inclusion by BC using the following criteria: 1) primary trial reports, 2) interventional trials, 3) human subjects, 4) phase 1 to phase 3, 5) trials of ixabepilone in monotherapy or combination therapy of 6) pre-licensure cancer indications. We excluded publications that were 1) secondary reports, 2) interim results, 3) meta-analyses or systematic reviews, 4) retrospective or observational studies, 5) laboratory studies of *ex vivo* human tissues,6) reviews, 7) letters, editorials, guidelines, interviews, abstract-only publications or poster presentations.

## Extraction

Our extraction template captured variables in the following domains: trial demographics, methods, hypotheses, trial design, measures of patient risk/burden and benefit (including adverse events, treatment duration, response and investigator conclusions). Criteria for extraction were pre-specified in a codebook (see appendix). All studies were extracted independently by two coders using Numbat software—an open-source meta-analysis management tool developed by BC.^9^ Disagreements were reconciled by discussion. When items were missing in reports, the corresponding author was contacted with 2 queries, with a response rate of 18%. Our codebook is available upon request.

Grade 3 or 4 serious adverse events (SAEs) defined by Common Terminology Criteria for Adverse Events (CTCAE)^10^ were scored conservatively, based on the highest number of events in a single CTCAE category per trial, since complete and per-patient SAE rates were rarely reported. Toxicities were only scored if they were defined as probably or definitely treatment-related in reports. In cases where the study did not mention whether events were treatment-related, these were not included.

The duration of treatment was defined to be the length of time a patient must be enrolled to collect the primary endpoint. When this number was not explicitly given, duration of treatment was imputed by the product of the median number of cycles and the period of each cycle.

Patient benefit was calculated in terms of the overall response rate (ORR) in individuals receiving ixabepilone on an intent-to-treat (ITT) basis. We used ORR because it is a widely used surrogate endpoint that allows us to compare effect sizes across trials of different indications and phases. Indications were divided into clinically meaningful categories identified in consultation with an oncologist.

A trial was considered to be a “positive” or “nonpositive” based on whether the study reached its pre-specified primary endpoint, and whether the regime was deemed tolerable by the authors. A trial was considered to be “uninformative” if any of the following conditions were met: 1) a nonpositive phase 2 trial was followed up with another trial in the identical patient population (duplication); 2) a positive phase 2 trial was not followed up with a confirmatory trial, either published or registered (no follow-up within 7 years of latest enrolment date); 3) a trial failed to reach 85% or more of its target sample due to factors other than futility or massive benefit (feasibility). We chose the second criterion on the assumption that phase 2 trials in cancer are geared towards being sensitive to possible new applications, and supporting a decision to advance a candidate for further testing. Failure to detect activity under these conditions would generally indicate an indication is unlikely to respond to a drug. Positive phase 2 trials that are not followed up are inadequate to guide clinical practice because they use surrogate endpoints only and/or are statistically underpowered; trials that fail to complete due to severe operational problems do not adequately address the hypothesis that justified their conduct.

## Analysis and Statistics

The rates of change of ORR and SAEs were calculated with R version 3.2.2.^11^ ORR and SAE rates were regressed against date of publication, weighted by sample size. We used a two-tailed test and defined *p*<0.05 as statistically significant. We did not adjust for statistical multiplicity. Pooling of responses and SAEs was done using the *meta* package for R,^12^ using a DerSimonian-Laird random effects model meta-analysis of proportions.

The differences in risk (death rate) and benefit (ORR) between both industry vs non-industry trials and phase 1 versus phase 2 trials in monotherapy were analyzed in R version 3.2.2^11^ using a weighted regression analysis. The outcome was regressed on a 0/1 indicator variable to identify industry trials or phase 1 versus phase 2 trials. The individual trial results were weighted by sample size, as the inverse variance of a rate estimate is directly proportional to sample size. We defined *p*<0.05 to be statistically significant.

## Results

### Study characteristics

Our search returned 62 primary publications of interventional trials testing ixabepilone in oncology in indications that were not approved at the time of trial initiation (see appendix). See Figure 1 for a PRISMA flow diagram and our appendix for details. We captured 39 monotherapy trials,^13-51^ and 23 combination therapy trials,^52-74^ spanning 14 different combination therapies. Our analyses included trials in indications that were not approved at the time the trial was initiated. Corresponding authors were based in 7 different countries. Properties of trials in our sample are described in Table 1.

**Table 1:**
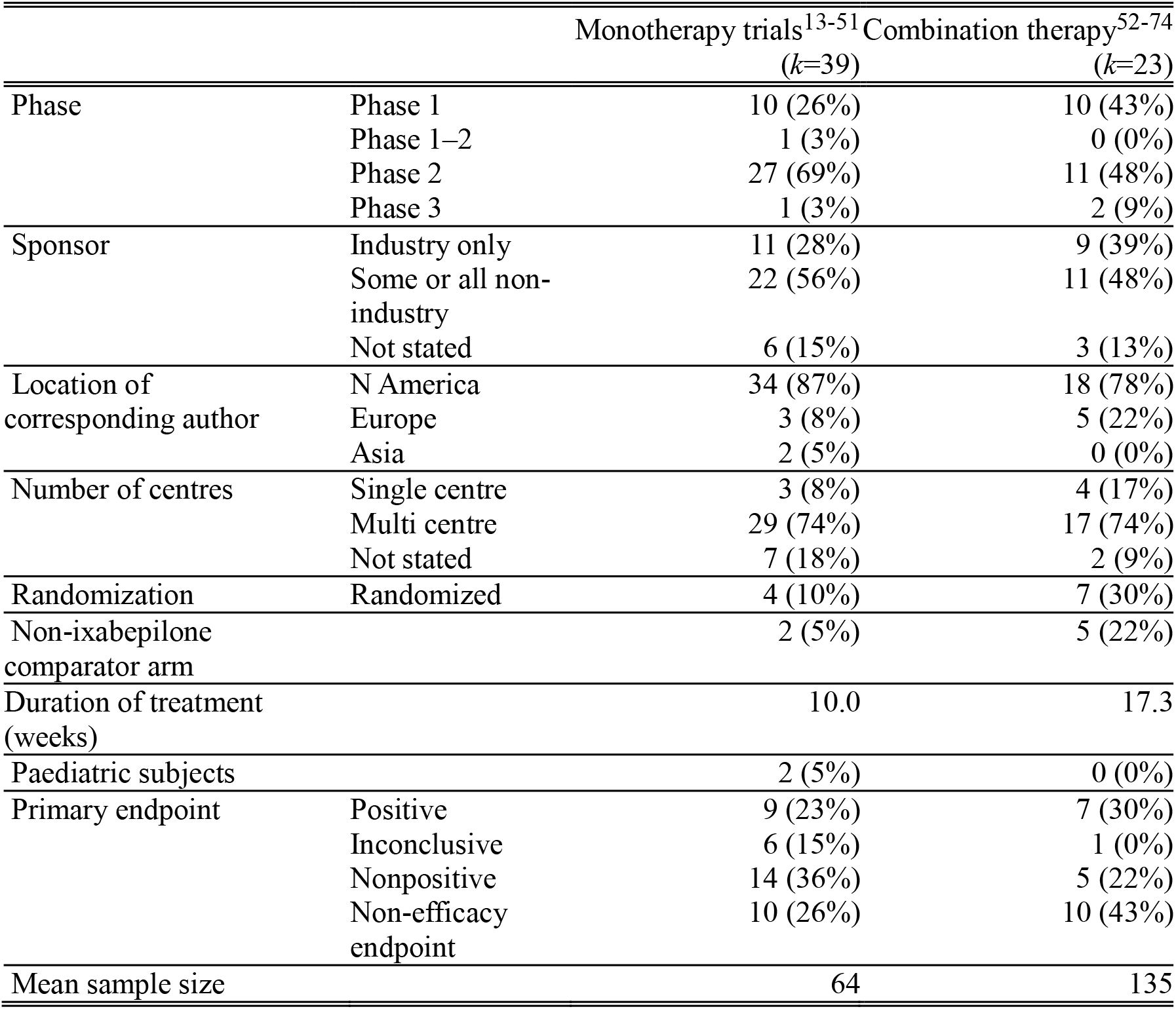
Properties of extracted trials in our sample

|  |  | Monotherapy trials<sup>13-51</sup><br>(*k*=39) | Combination therapy<sup>52-74</sup><br>(*k*=23) |
| --- | --- | --- | --- |
| Phase | Phase 1 | 10 (26%) | 10 (43%) |
|  | Phase 1–2 | 1 (3%) | 0 (0%) |
|  | Phase 2 | 27 (69%) | 11 (48%) |
|  | Phase 3 | 1 (3%) | 2 (9%) |
| Sponsor | Industry only | 11 (28%) | 9 (39%) |
|  | Some or all non-industry | 22 (56%) | 11 (48%) |
|  | Not stated | 6 (15%) | 3 (13%) |
| Location of corresponding author | N America | 34 (87%) | 18 (78%) |
|  | Europe | 3 (8%) | 5 (22%) |
|  | Asia | 2 (5%) | 0 (0%) |
| Number of centres | Single centre | 3 (8%) | 4 (17%) |
|  | Multi centre | 29 (74%) | 17 (74%) |
|  | Not stated | 7 (18%) | 2 (9%) |
| Randomization | Randomized | 4 (10%) | 7 (30%) |
| Non-ixabepilone comparator arm |  | 2 (5%) | 5 (22%) |
| Duration of treatment (weeks) |  | 10.0 | 17.3 |
| Paediatric subjects |  | 2 (5%) | 0 (0%) |
| Primary endpoint | Positive | 9 (23%) | 7 (30%) |
|  | Inconclusive | 6 (15%) | 1 (0%) |
|  | Nonpositive | 14 (36%) | 5 (22%) |
|  | Non-efficacy endpoint | 10 (26%) | 10 (43%) |
| Mean sample size |  | 64 | 135 |

**Figure 1:**
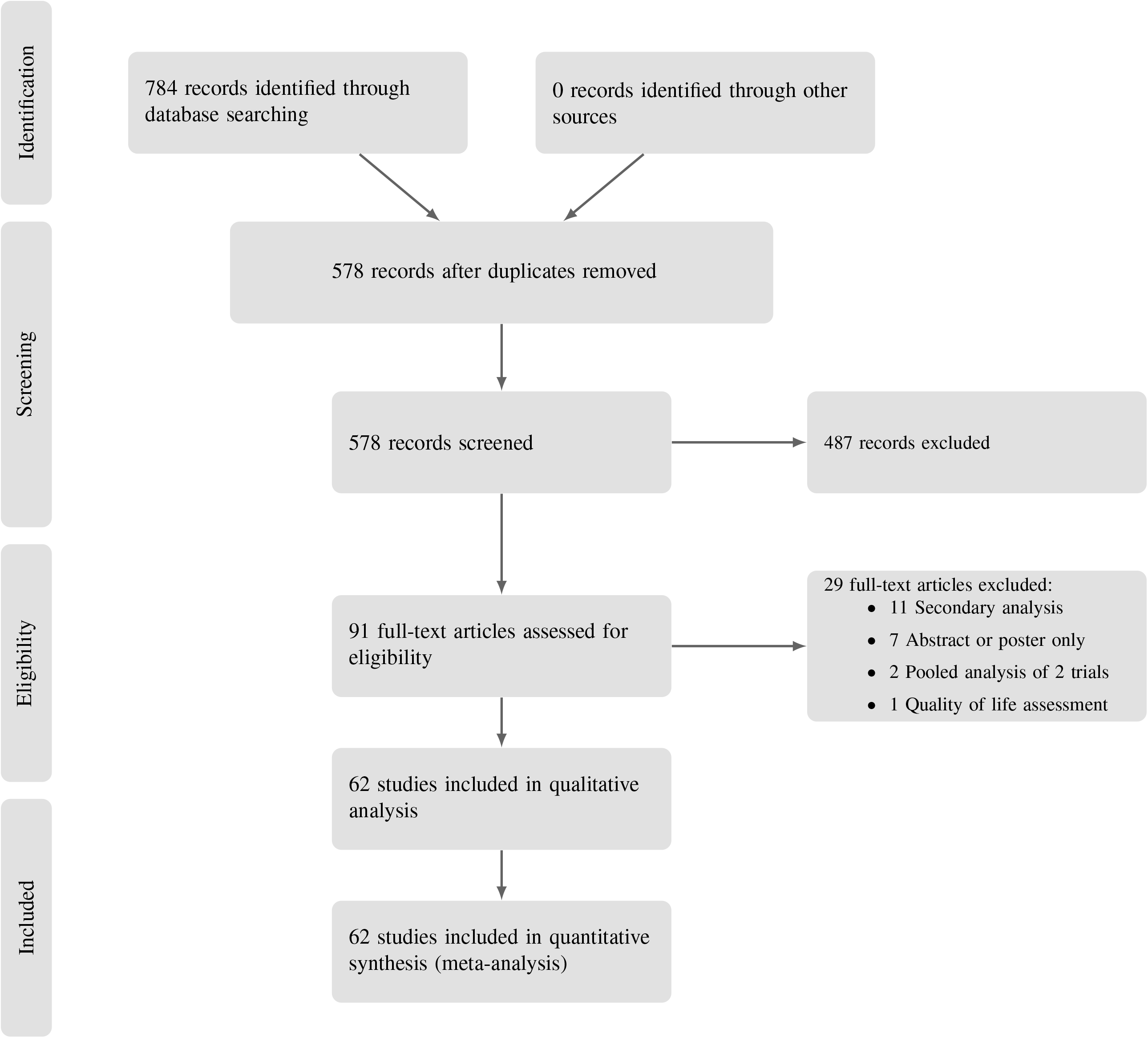
PRISMA flow diagram

Using the U.S. National Library of Medicine (NLM) clinical trials registry, ClinicalTrials.gov, we estimate an additional 20 monotherapy or combination therapy trials were completed but not published as full-text publications within 4 years of closure (see appendix). This included another 5 malignancies. Another 35 trials identified in the NLM registry are either ongoing as of this writing, or may have closed too recently for publication.

### Total patient benefit and burden

In total, 830 patients receiving ixabepilone experienced objective tumour response (16%, 95% CI 12.5%–20.1%), and 74 died from drug-related toxicites (2.2%, 95% CI 1.6%–2.9%). We estimate a minimum of 1276 experienced grade 3–4 drug-related severe adverse events (34.4%, 95% CI 29.9%–39.2%).

The 39 trials included in the ixabepilone monotherapy analysis involved 2504 patients and 546 patient-years of participation over a period of 13 years. Seven different doses of ixabepilone monotherapy were tested in the translation trajectory of ixabepilone, using 7 schedules and testing 15 different malignancies.

In monotherapy, 241 patients receiving ixabepilone experienced objective tumour response (11.1%, 95% CI 8.6%–14.3%); 29 died from drug-related toxicities (2.2%, 95% CI 1.6%–3.0%); a minimum of 667 experienced grade 3–4 drug-related toxicities (31.3%, 95% CI 25.7%–37.5%). These figures compare with an ORR of 18.3% (95% CI 11.9%–26.1%) and a SAE rate of 42.9% (95% CI 34.3%–51.5%) in the CTCAE-defined category (leukopenia) with the highest event rate in the monotherapy trial cited in the FDA approval.^33^

In combination therapy, 3111 patients and 1075 patient-years of involvement over 13 years in 9 different countries, testing 14 different combination therapies were sampled. In total, 589 patients experienced objective tumour response (29.6%, 95% CI 22.3%–38.1%), and 45 patients died from treatment-related toxicity (1.9%, 95% CI 1.1%–3.3%). There was a minimum of 636 grade 3–4 drug-related toxicities (39.4%, 95% CI 32.4%–46.8%). These figures compare with an ORR of 34.7% (95% CI 29.9%–39.7%) and a severe adverse event rate of 39.1% (95% CI 35.6%–42.6%) in the single CTCAE-defined category (neutropenia) with the highest event rate in the pivotal combination therapy trial leading to FDA approval.^55^

### Clinical development trajectory

To visualize the progression of testing during ixabepilone development, we prepared Accumulation of Evidence and Research Organization (AERO) diagrams of all monotherapy and combination therapy trials (Figures 2 and 3).^75^ Though the first indication tested (colorectal cancer) proved nonresponsive, the second (breast cancer) responded and was quickly licensed in the U.S. Subsequent to licensure, further breast cancer trials tested ixabepilone in and against different combinations, and with different biomarkers. By 2010, drug developers appeared to have discontinued testing new indications for ixabepilone monotherapy. The latest unpublished novel indication pursued according to trial registry records was urethral and bladder cancer, which closed due to accrual failure in 2007.

**Figure 2:**
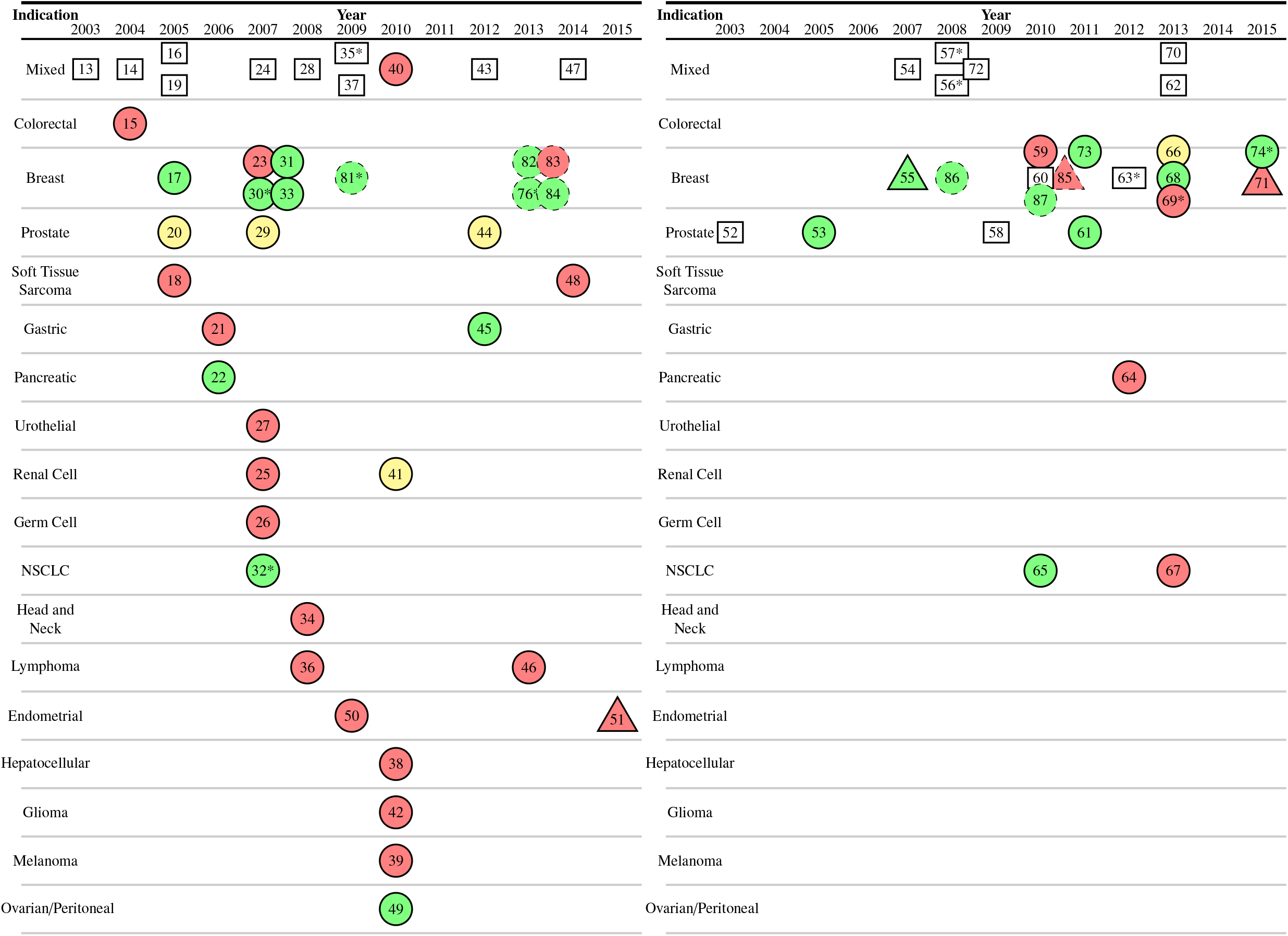
Ixabepilone monotherapy (left) and combination therapy (right) AERO graph. Trials arranged by publication year horizontally; stratified by indication vertically. Rectangular nodes are phase 1 trials. Circular nodes are phase 2 trials. Triangular nodes are phase 3 trials. Green nodes indicate positive primary endpoint and acceptable toxicity. Yellow indicates inconclusive primary endpoint. Red indicates nonpositive primary endpoint or unacceptable toxicity. White indicates non-efficacy endpoint. Dashed borders indicate trials launched in an indication that had already received FDA licensure (these were not included in our analysis). Asterisks indicate European corresponding author.

**Figure 3:**
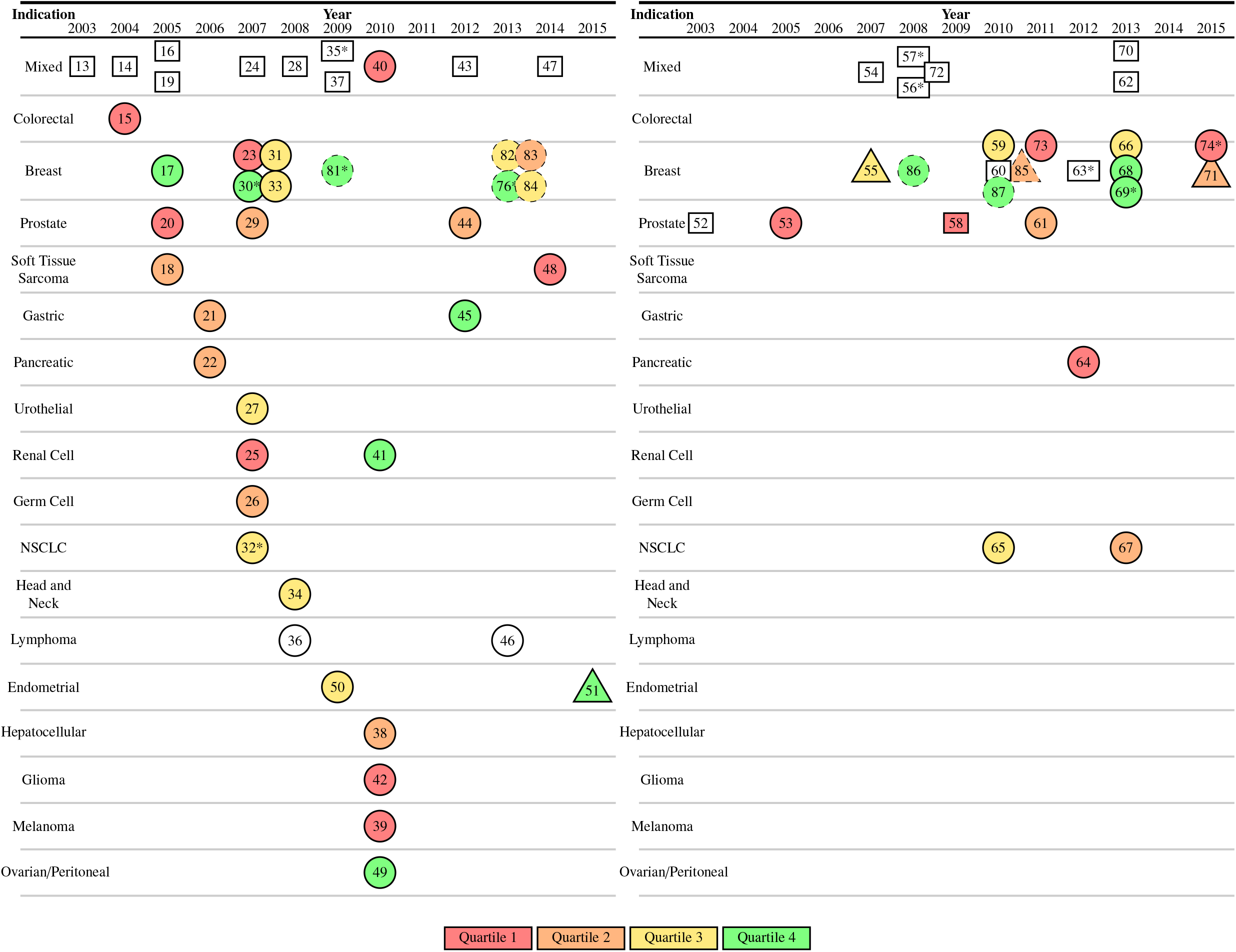
Ixabepilone monotherapy (left) and combination therapy (right) AERO graph. Trials arranged by publication year horizontally; stratified by indication vertically. Rectangular nodes are phase 1 trials. Circular nodes are phase 2 trials. Triangular nodes are phase 3 trials. Nodes are coloured by ORR divided into quartiles, with separate quartiles for mono- and combination therapy. White colouration indicates non-efficacy primary endpoint or non-RECIST response measurement. Dashed borders indicate trials launched in an indication that had already received FDA licensure (these were not included in our analysis). Asterisks indicate European corresponding author.

Figures 2 and 3 (right panels) show the exploration of combination therapy in 4 different indications. Ixabepilone was first licensed by the FDA in 2007 for treatment of metastatic or locally advanced breast cancer in combination with capecitabine after failure of an anthracycline and a taxane, and in monotherapy after failure of an anthracycline, a taxane and capecitabine.^5^ Ixabepilone was rejected by the European Medicines Agency (EMA) for licensure in 2008.^8^ Total patient burden in relation to milestones in the development of ixabepilone is depicted in Figure 4. Dosing and scheduling reflected on the FDA label, as well as responding malignancies, were discovered with very little patient burden (10 grade 3–4 events; less than 1% of the grade 3–4 events in the total portfolio; 8 patient-years of involvement; less than 1% of 1598 patient-years in trials of non-approved indications). The sharpest incline in cumulative treatment-related deaths occurred after efficacy in breast cancer was established sufficiently for FDA approval; this sharp increase represented 29 trials in 15 indications. Most (68%) of the grade 3–4 events and 89% of deaths in the total portfolio of non-approved indications occurred after the first and only responding indication was identified.

**Figure 4:**
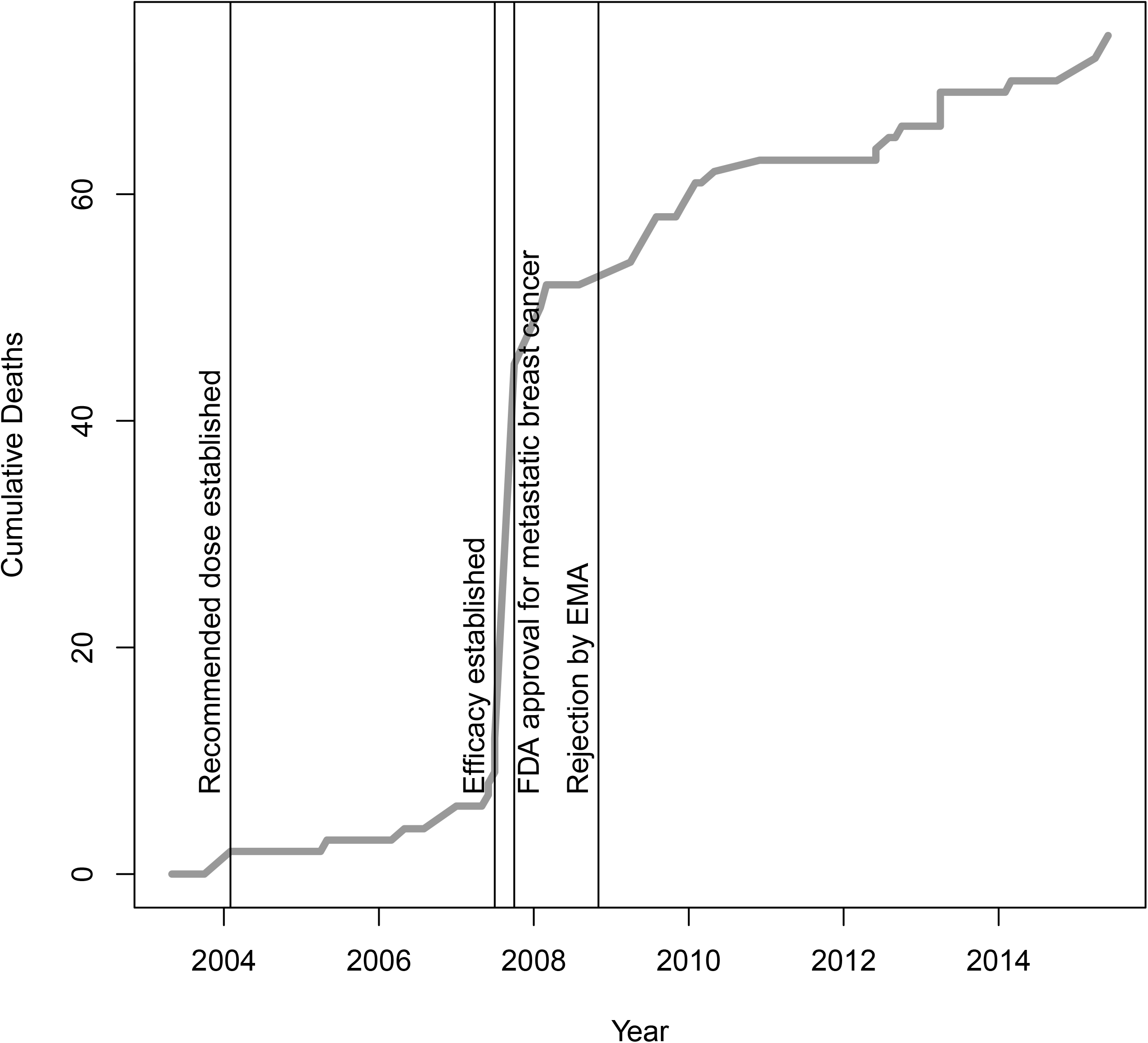
Cumulative treatment-related deaths in trials of ixabepilone in indications not approved by the FDA at time of patient enrolment for key milestones: recommended dose established (2004), the publication of the phase 2 trial in which effectiveness was first established,^31^ the FDA approval of ixabepilone for metastatic breast cancer in October 2007,^5^ rejection of ixabepilone by the EMA in November 2008.^8^ The total number of deaths in the whole portfolio is charted against time.

In our AERO diagrams, unfavorable licensing outcomes in Europe appeared to affect the geography of clinical testing. Before EMA’s rejection, 10 trials were reported as having been conducted in Europe (these were published after the EMA rejection; see Figures 2–3). After, there were no further trials that enrolled patients and that included corresponding authors in Europe. The remainder of the trials pursuing new licenses were performed by corresponding authors based in North America.

### Trends for Risk, Benefit and Success

We plotted the evolution of risk and benefit across the trajectory of ixabepilone development including mono- or combination therapy trials of indications that were not approved by the FDA (Figure 5). We did not observe a significant departure from a zero slope in drug-related adverse event rates (*p*=0.57). There was a significant departure from a zero slope in ORR (*p*=0.001), but this reflects an increase in the exploration of combination therapy, as there was no departure from a zero slope for studies in monotherapy only (not shown, *p*=0.9).

**Figure 5:**
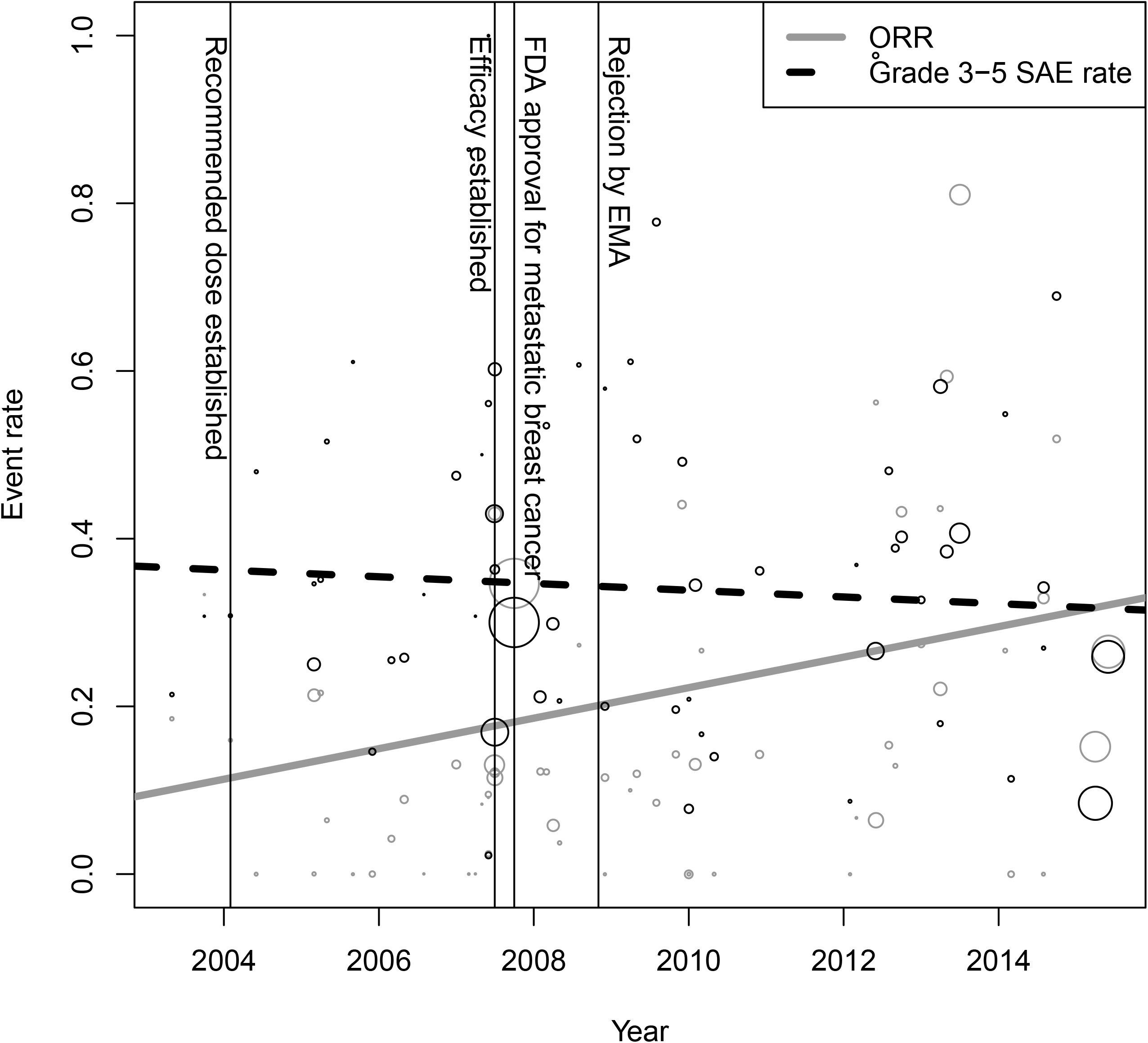
Risk and benefit trends for studies in non-approved indications in both monotherapy and combination therapy. All events were classified by report authors as probably or definitely drug related.

Industry-funded studies in indications that had not received FDA licensure at the time of trial initiation had a higher ORR than non-industry-funded studies (33.9% vs 20.7%, *p*=0.005), but the rate of treatment-related death for industry-funded studies was not significantly different from the rate of non-industry-funded studies (46 out of 1790 patient-subjects, 4.7% vs 23 out of 1856 patient-subjects, 2.3%, *p*=0.058).

For monotherapy trials in non-FDA-approved indications, phase 2 trials had a higher response rate, but not significantly so (15.7% vs 10.0%, *p*=0.2). Rates of treatment-related deaths were also not significantly different (2.3% vs 3.0%, *p*=0.9).

A total of uninformative 11 trials were found, 7 for lack of appropriate follow-up, 4 for accrual failure and 0 for duplicity.^22,26,32,46,49,53,61,65,66,74,76^ These trials represent 792 patients and 226 patient-years of involvement in which there was a minimum of 208 grade 3–4 severe adverse events (27.8%, 95% CI 22.4%–33.9%) and 7 deaths occurred (1.4%, 95% CI 0.8%– 2.7%). Among these patients, 175 experienced objective tumour response (24%, 95% CI 16.7%– 33.3%). The grade 3–4 severe adverse events and patient-years of involvement among the uninformative trials represent 21% and 26% of the portfolio total, respectively.

The phase 3 trial of endometrial cancer did not meet our criteria for inclusion in our set of uninformative trials, however it is worth noting because it was launched after a prior phase 2 failure in the same indication and represents 496 patients receiving ixabepilone monotherapy, in which two patients on the ixabepilone arm died from study drug toxicity.^51^

## Discussion

In a previous characterization of risk, benefit and success rates for the development of the targeted drug sunitinib, we observed worsening of risk/benefit, exhaustive testing of different indications, and duplicative nonpositive trials, suggestive of excess burden.^3^ Our study of the trajectory of development for ixabepilone—a non-targeted drug—offers parallels and contrasts.

As with sunitinib, ixabepilone developers very quickly discovered conditions for the first FDA label and with little patient burden. For instance, the second of numerous malignancies tested in trials of both sunitinib and ixabepilone proved to be the most responsive. As also observed with sunitinib, most of the burden in testing ixabepilone against non-approved indications occurred after the first licensed indication, not before. Similarly to sunitinib, returns of further indication and combination exploration for ixabepilone were steeply and vastly diminished.

We also found that benefit for phase 2 trials exploring new indications was only slightly better than benefit levels observed in phase 1 studies (15.7% vs 10.0%, respectively, *p*=0.2). Risk of treatment-related death was not significantly different (2.3% vs 3.0%, respectively, *p*=0.9). A pattern of worsened risk/benefit in phase 2 studies was observed for sunitinib.

In contrast with the development of sunitinib, we did not observe a worsening of risk/benefit as the clinical development portfolio of ixabepilone trials matured, nor did we observe as much perseverance and seeming duplication in testing. Several indications that failed to produce significant signal of activity in initial phase 2 trials were followed up in subsequent investigations. Ixabepilone developers were especially persistent in pursuing four indications— prostate cancer, non-small cell lung cancer, lymphoma and pancreatic cancer.

For example, prostate cancer was tested in 3 monotherapy and 4 combination therapy studies. The combination studies met their primary endpoints with caveats regarding toxicity and the need for a decisive phase 3 trial in this indication has been highlighted as early as 2007.^77^ There is still no published phase 3 trial, and as of this writing, no U.S. registered phase 3 trial in prostate cancer. This represents 106 patient-years expended in developing ixabeplione for prostate cancer that have gone unredeemed insofar as promising findings have not been followed up with decisive testing. One monotherapy and two combination therapy trials were pursued for non-small cell lung cancer. Lymphoma showed promising activity in an early phase 2 monotherapy study.^36^ This was confirmed in a later phase 2 trial, but the authors declared ixabepilone unlikely to be useful for this indication because of the availability of “numerous” promising alternatives (e.g. rituximab, which became available soon after this study was started).^46^ A nonpositive phase 2 trial of soft tissue sarcoma included a subgroup of 8 responding patients with leiomyosarcoma.^18^ This was followed by a trial in uterine leiomyosarcoma; the study was nonpositive and had lower response rate than the first trial.^48^ Endometrial cancer was explored in a phase 2 trial published in 2009. This trial also failed its primary endpoint and was described by the authors as having only “modest activity of limited duration.”^50^ Nevertheless, a phase 3 trial was initiated after the phase 2 was published, in which there were 2 treatment-related deaths and 164 drug-related life threatening (grade 3–4) toxicities.^51^ The National Comprehensive Cancer Network has made a Category 2B (lower-level evidence) recommendation of second-line ixabepilone monotherapy for recurrent, metastatic, or high-risk endometrial carcinoma.^7^ Our analysis of response rates and severe adverse events for patient-subjects enrolled in trials of non-approved indications of ixabepilone did not reveal a clear trend in improving or worsening risk/benefit as the translation trajectory matured. This marks a contrast with sunitinib.

As illustrated in Figure 4, the major burdens for patients enrolling in trials of ixabepilone were incurred between the initial evidence of efficacy and the rejection by the EMA in November 2008. After this point, research activity for ixabepilone cooled, and the last new indication explored in a phase 2 trial of ixabepilone was in 2010.^42^ Some investigators blamed this regulatory action for difficulty in recruiting patients.^76^

The study of ixabepilone provided an occasion to explore how trial activities change in response to regulatory decisions. Trial activities in Europe were discontinued after an unfavorable licensing decision by the EMA for breast cancer. However, after ixabepilone was approved in the U.S., drug developers launched a minimum of 11 trials and 5 breast cancer combination trials. If licensing were being sought for these indications or combinations, these same trials might have been pursued in Europe as well. One possible explanation for this geographical shift is that ixabepilone’s developers, calculated that the probability of demonstrating decisive benefit superior to breast cancer for other indications or drug combinations was marginal. However, such an explanation invites questions of why so much testing was pursued in other jurisdictions. Another potential explanation is that some trial activities that occur after an initial license are primarily aimed at encouraging off-label drug use or uptake into guidelines, regardless of licensure. Participation in these “seeding” trials can increase the probability that physicians write prescriptions for a drug,^78^ and drug companies have sometimes deliberately used trials to promote off-label drug use.^79^ As many as 50% of cancer drug prescriptions in Europe and the U.S. are off-label.^80^ That ixabepilone was not approved in Europe foreclosed the prospect of off-label use there. Hence, drug developers may have instead shifted activities to a jurisidiction where off label use and/or uptake in guidelines was possible. Nevertheless, this explanation would be very difficult to test absent company documents that are inaccessible.

Our analysis has several limitations. First, this study relied on published full reports of trials. Gaps in the literature (unpublished trials, abstract-only publications, etc.) limit our ability to capture all events and patient responses. However, any publication bias in the ixabepilone translation trajectory would be unlikely to show a more favourable pattern of risk/benefit. Severe adverse event rates were not reported consistently. In 30% of studies, serious adverse events were only reported above a certain threshold; as a result, we have underestimated risk. Second, we have used response rates as a surrogate for benefit. The relationship between response rate and clinical benefit is not always clear—as illustrated by the fact that many indications showed response rates similar to those in trials leading to FDA approval.

In short, based on these findings and the contrasts with sunitinib, we offer the following hypotheses about risk/benefit for development of novel cancer drugs: First, cancer drug developers are very effective at using preclinical and early phase clinical trial evidence to identify the most promising conditions for licensure of a new drug. Few trials, patients-years and toxicities are needed to achieve a level of activity that can lead to licensure. Second, much of the risk and burden in drug development occurs after products receive licensure and drug developers attempt to extend the label. Gains in knowledge about the properties of novel drugs do not seem to lead to any diminution of event rates, or marked improvements in success, and gains in clinically useful knowledge diminish sharply after initial licensure. Third, once a drug receives an initial approval in one jurisdiction, drug developers will concentrate their research activities in the same jurisdiction.

## Data Availability

Data attached as supplementary materials

## Acknowledgement

This work was funded by CIHR (EOG111391).

